# Restricting carbohydrates and calories in the treatment of type 2 diabetes: a systematic review of the effectiveness of ‘low carbohydrate’ interventions with differing energy levels

**DOI:** 10.1101/2021.05.07.21256843

**Authors:** Anna P Nicholas, Adrian Soto Mota, Helen Lambert, Adam L Collins

## Abstract

There are two proven dietary approaches to shift type 2 diabetes (T2D) into remission: low energy diets (LED) and low carbohydrate diets (LCD). These approaches differ in their rationale and application yet both involve carbohydrate restriction, either as an explicit goal or as a consequence of reducing overall energy intake. The aims of this systematic review were to identify, characterise and compare existing clinical trials that utilised ‘low carbohydrate’ interventions with differing energy intakes. Electronic databases CENTRAL, CINAHL, Embase, MEDLINE and Scopus were searched to identify controlled clinical trials in adults with T2D involving low carbohydrate intake (defined as <130g carbohydrate/day) and reporting weight and glycemic outcomes. The initial database search yielded 809 results, of which 18 studies met the inclusion criteria. 12/18 studies utilised low carbohydrate diets with moderate or unrestricted energy intake. Six trials utilised low energy diets (<1200kcal/day), with all except one incorporating meal-replacements as part of a commercial weight loss programme. Interventions using both restricted and unrestricted (*ad libitum*) energy intakes produced clinically significant weight loss and reduction in HbA1c at study end-points. Trials that restricted energy intake were not superior to those that allowed *ad libitum* low carbohydrate feeding at 12 and 24 months. An association was observed across studies between average weight loss and reduction in HbA1c, which strengthened with trial length, indicating that sustained weight loss is key to T2D remission. Further research is needed to specifically ascertain the weight-independent effects of carbohydrate restriction on glycemic control in T2D.

**Graphical abstract:** 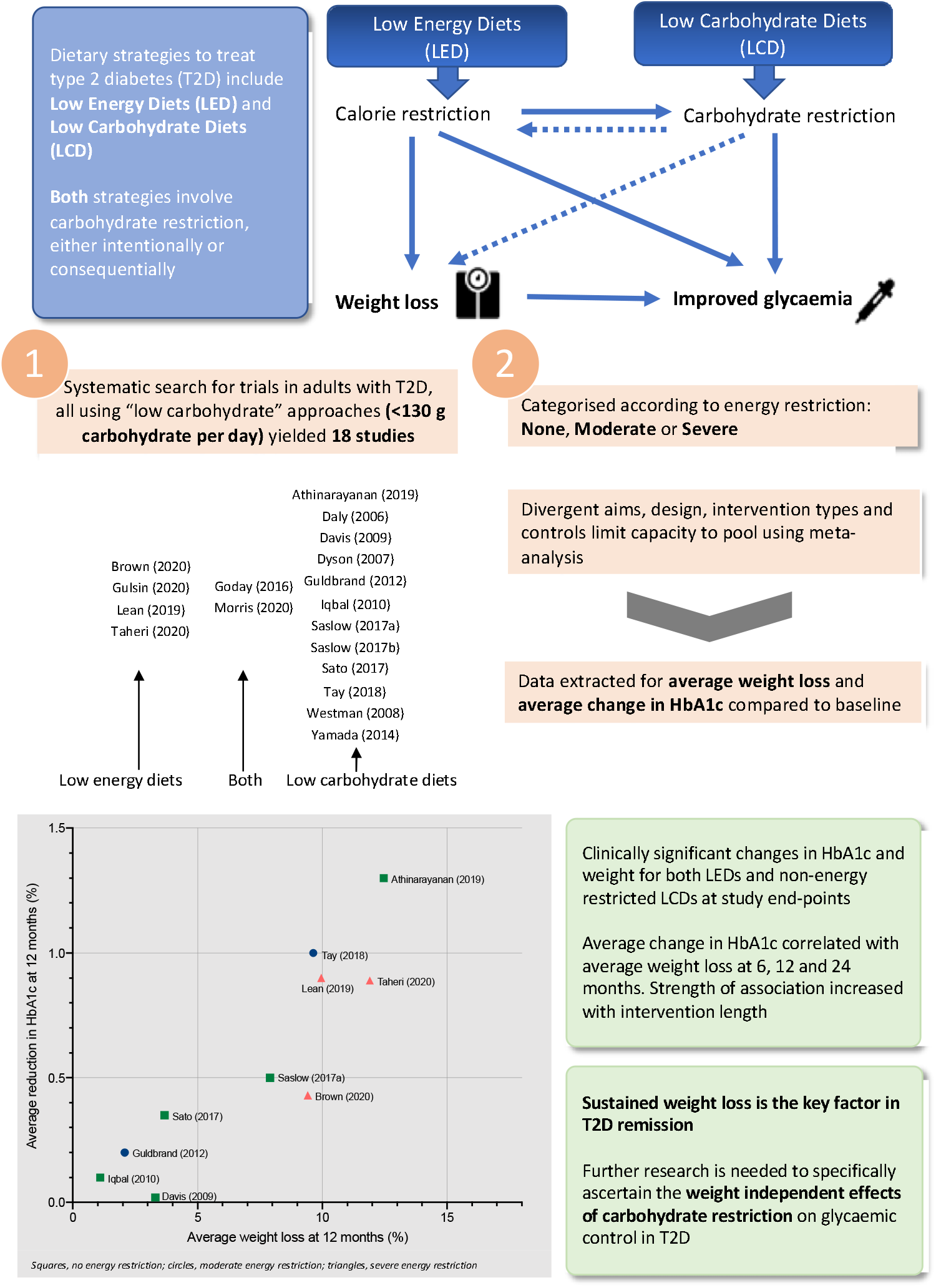

## Introduction

The prevalence of type 2 diabetes (T2D) has reached epidemic proportions and is a major global concern. According to the World Health Organisation (WHO), more than 422 million people have diabetes worldwide, representing a global prevalence among adults of 8.5% (1). In the UK alone, over 3.9 million people are diagnosed with diabetes, 90% of which have T2D, and this figure is anticipated to rise to more than 5 million by 2025 (2). T2D is a major risk factor for other health conditions including cardiovascular disease (CVD), kidney failure, neuropathy and blindness (1). It has also recently emerged as a significant risk factor for Covid-19 (3).

T2D used to be considered a chronic progressive disease typically managed by escalating pharmacotherapy to maintain normoglycemia and mitigate disease complications. However, the paradigm of treatment is changing with recognition that T2D can be put into remission (4). This remission, it seems, can occur up to a point, beyond which the pancreatic beta-cells are unable to recover (5). Definitions for remission vary but it is generally defined as achieving fasting plasma glucose (FPG) <7mol/L or glycated haemoglobin (HbA1c) <6.5% (48mmol/mol) for a sustained period of time (2 to 12 months) and in the absence of antiglycemic medications (4).

There are currently two proven non-surgical ways to achieve T2D remission: low energy diets (LED) and low carbohydrate diets (LCD) (6–8). These two approaches focus on operating different metabolic levers: energy restriction and carbohydrate restriction. Given both factors are interlinked (Figure 1), it is not clear which is driving T2D remission and hence which offers the most effective interventional approach.

**Figure 1:**
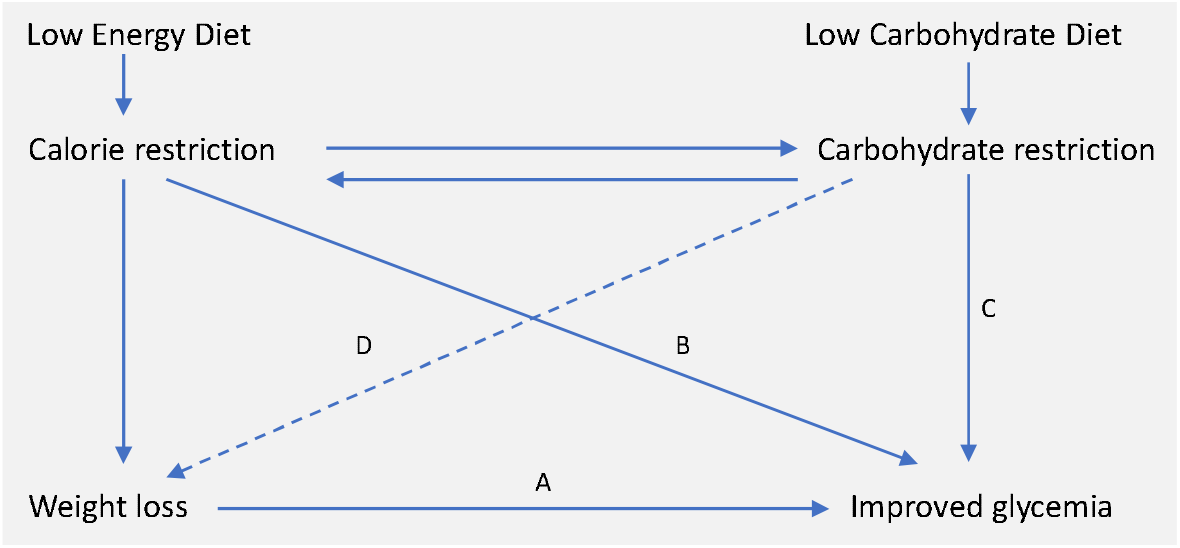
Interrelationship between energy restriction, weight loss and carbohydrate restriction in improved glycemic control. Carbohydrate and energy restriction are interrelated. A) In obese individuals with T2D, weight loss is associated with improved glycemic control (9). This is in accordance with the twin cycle hypothesis, whose central tenet is that excess lipids within the liver and the pancreas drive T2D pathogenesis (10). B) In studies of low energy feeding, glycemia improves within days of energy restriction, before significant weight loss has occurred (11). C) Carbohydrate restriction improves glycemia by reducing postprandial glucose rises. While failed repression of gluconeogenesis and glycogenolysis are major causes of hyperglycemia (12), dietary carbohydrate intake is the largest driver of postprandial glucose rises. D) Carbohydrate restriction is also associated with weight loss. This may occur as a function of spontaneous energy restriction or there may be independent effects arising from reduced insulin secretion. Whether or not carbohydrate restriction has independent effects on body weight remains a matter of contentious debate (hence depicted as dashed line) (13,14).

LEDs restrict energy intake to induce rapid weight loss (15). They characteristically provide between 800 and 1200 kcal/day utilising either total diet replacement (TDR) or some inclusion of conventional foods as a partial diet replacement (16). LEDs have gained attention for their use in diabetes management after the 2011 Counterpoint study demonstrated reversal of abnormalities underlying T2D using an 8 week 600kcal/day diet (17). In 2018, the DiRECT trial demonstrated that an intensive weight management programme using TDR could achieve T2D remission in 46% of participants after one year (7).

LCDs specifically aim to restrict carbohydrate intake, either as a percentage of total energy or as an absolute intake (grams per day). Definitions of what constitutes ‘low carbohydrate’ have been inconsistent over time and between studies, but the definitions proposed by Feinman et al (18) are becoming more widely accepted. Specifically, this defines low carbohydrate as less than 130g CHO/day and very low carbohydrate ketogenic diets as less than 50g/day.

There is currently much interest in the application of LCDs to T2D (19). Over the past five years, 10 meta-analyses, based on nearly 50 randomised controlled trials (RCTs), have aimed to address the question of whether diets low in carbohydrates produce greater improvements in weight and glycemic control compared to higher carbohydrate control diets (20–29). The majority of these meta-analyses have found a beneficial effect from carbohydrate restriction (20–26), and none have favoured higher carbohydrate comparators, although several studies have found no difference between diets (27–29).

The table below highlights the similarities and differences between LCDs and LEDs. It is clear that, despite their different aims, carbohydrate restriction is common in both: LCDs restrict carbohydrates as an explicit goal, whereas LEDs restrict carbohydrates as a consequence of achieving low energy intake.

### Study aims

Systematic reviews of carbohydrate restriction interventions to date have focussed on traditional LCDs but have not included LEDs that are also ‘low carbohydrate’ in absolute terms. Moreover, most have also used a range of definitions for LCDs, from <25% to <50% TE (20,27). This review takes an alternative approach to the existing evidence-base by recognising the commonality between these two approaches “clamped” by carbohydrate intake. Specifically, it aims to review, characterise and compare the clinical trials that have used low carbohydrate (<130g/day) approaches with different levels of energy intake.

## Methods

### Data sources and searches

The present systematic review was performed with reference to the Cochrane Handbook for Systematic Reviews of Interventions (30) and reported in accordance with the Preferred Reporting Items for Systematic Reviews and Meta-Analyses (PRISMA) statement (31). A protocol was registered with PROSPERO in advance (CRD42020197257) (32).

An electronic search was performed using the databases Medline, EMBASE, CINAHL, Scopus and Cochrane Central Register of Controlled Trials (CENTRAL). The search was performed on 7^th^ July 2020 and no date limits were applied. Search terms included keywords and subject headings related to T2D, low energy or low carbohydrate diets, glycemic outcomes and clinical trials (see Supplemental Materials). A manual search of reference lists of key systematic reviews and reports was also conducted to identify any additional relevant studies. Search results were independently reviewed by A.N and A.S.M and any conflicts over inclusion were resolved by discussion.

### Study selection

Studies were eligible for inclusion if they were controlled trials including adults diagnosed with T2D, involving a low carbohydrate diet (defined as <130g/day or <26% of total energy) and reporting change in weight and glycemic control. Non-randomised trials were eligible to allow inclusion of trials in more ecologically valid settings, such as those utilising very low energy weight loss diets. Control diets that stipulated any other type of dietary intervention such as low fat, ‘healthy eating’ and Mediterranean, or usual diabetes care were permitted. All countries and languages were eligible. For full inclusion and exclusion criteria, see Table 2.

**Table 1:** Generalised similarities and differences between Low Carbohydrate Diets and Low Energy Diets.

|  | Low Energy Diets | Low Carbohydrate Diets |
| --- | --- | --- |
| Meal format | Usually meal replacements soups, shakes and bars | Usually food-based |
| Energy restriction | Energy restricted to $\leq 1200$ kcal/day | Variable. Some allow *ad libitum* feeding to satiety; others include moderate energy restriction ($>1200$ kcal/day) |
| Carbohydrate restriction | Variable carbohydrate content but usually at least 50g CHO/day | Restricted to $<130$g CHO/day; VLCKD restricted to 20-50g CHO/day |
| Duration | Necessarily restricted to short periods of up to 3 to 5 months | No limit on duration |

**Table 2:** Study inclusion and exclusion criteria. CCTs, controlled clinical trials; CHO, carbohydrate; CVD, cardiovascular disease; RCTs, randomised controlled trials; T1D, type 1 diabetes; T2D, type 2 diabetes; TE, total energy

| Inclusion criteria | Exclusion criteria |
| --- | --- |
| RCTs and CCTs using low CHO diets or very low energy diets with <130g/day or <26% TE from CHO in adults (≥18 y) with T2D | Studies that included people with other chronic diseases (except hypertension or CVD) or taking systemic corticosteroids, or had any progressive disease requiring hospital care |
| Control is any other type of dietary intervention or usual care | Studies involving participants undergoing surgery |
| Data from crossover trials with washout periods of ≥4 weeks between interventions. In absence of adequate wash-out period, data from these trials only included if able to extract relevant data for 1st phase | Studies of T1D, prediabetes or gestational diabetes |
| Studies of individuals with and without T2D only if subgroup analysis available for participants with T2D | Studies with enteral or parenteral feeds |
| Studies reporting change in weight and markers of glucose control | Participants aged <18 y or pregnant/lactating women |
| Provided macronutrient goals/CHO goals as TE or g/day | Treatment diet poorly defined; unclear whether low CHO criteria were met |
|  | Studies involving intermittent fasting protocols |
|  | Study duration <1 week |
|  | Studies based on medication co-intervention not applied to all groups |

### Data extraction and quality assessment

Data were extracted by A.N. Data items included: study characteristics, participant characteristics, details of the intervention (including prescribed and reported energy and macronutrient composition), dietary adherence and outcome data for HbA1c and weight loss at 3, 6, 12, 24 months (where available) and study-end points. HbA1c only was collected as it is the most widely used marker of T2D remission and medication changes and quality of life were not reported due to lack of consistency across studies. These represent minor deviations from the protocol submitted to PROSPERO. The mean percentage weight loss from baseline and mean absolute reduction in HbA1c were calculated for intervention arms. Absolute rather than relative change in HbA1c was used since therapeutic goals are based on a threshold value and not relative reduction (33). Risk of Bias was assessed using the Cochrane Risk of Bias tool (34).

### Data synthesis and analysis

A narrative synthesis was undertaken to explore characteristics of the included studies. HbA1c and weight loss outcome data were compared between intervention and control arms within studies at the longest duration time-point.

In order to compare weight and HbA1c outcomes between studies, percentage weight loss and HbA1c change at study end-point and at specific time points (3, 6, 12 and 24 months) were plotted graphically in scatter plots. Meta-analysis was not considered appropriate due to high clinical heterogeneity between studies. The association between average weight loss and HbA1c change was examined using correlation analysis and computation of R-squared values in Prism 8 for OS X Version 8.4.3 (35).

## Results

### Search results

Figure 2 shows the selection of studies, in accordance with the PRISMA guidelines (31). The initial database search yielded 809 studies, of which 223 were duplicates. Following title and abstract screening, 91 studies were retrieved for full text screening. A total of 18 studies met the inclusion criteria.

**Figure 2:**
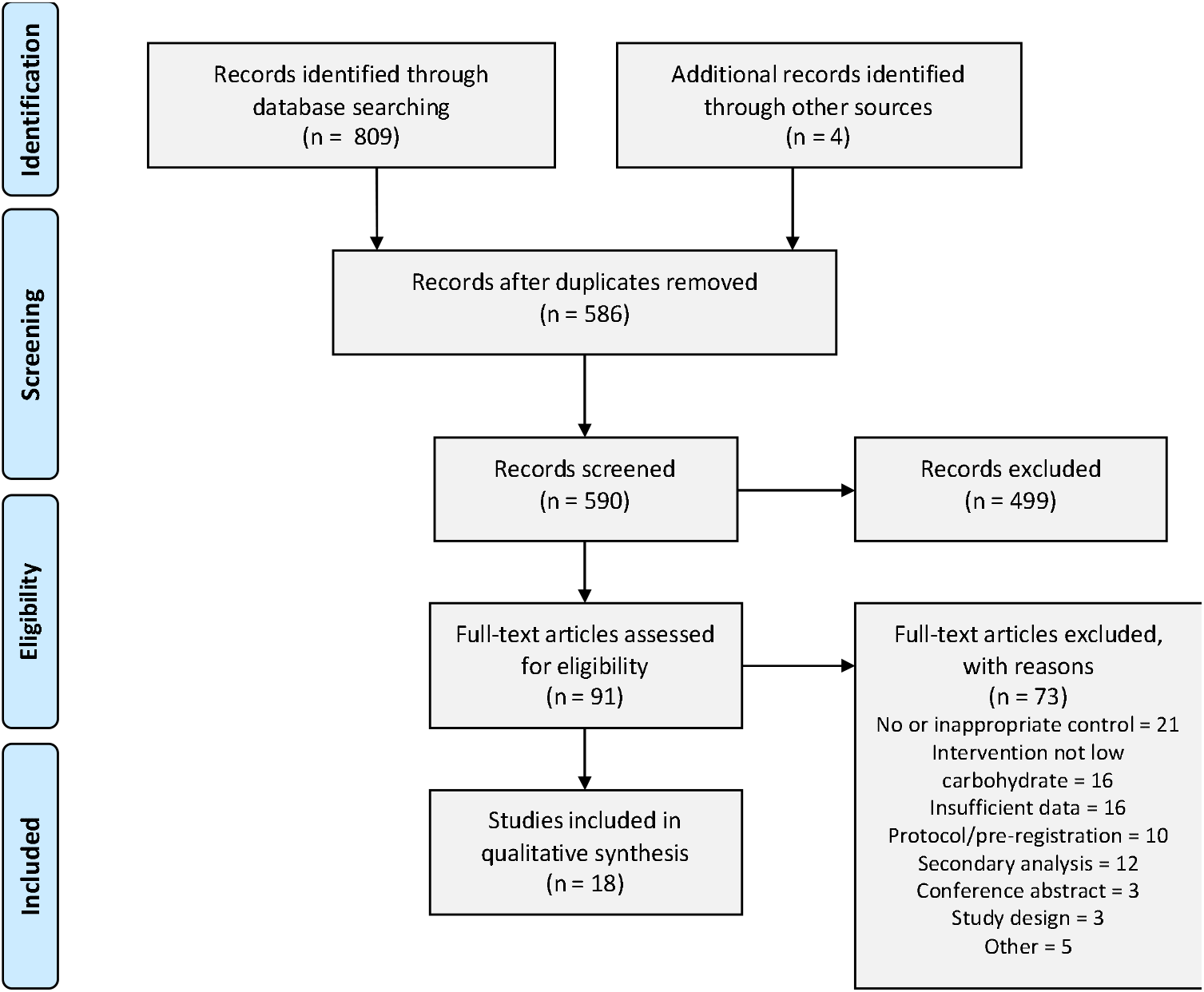
Study screening and selection.

### Study characteristics

This review yielded a highly heterogenous set of studies that fulfilled criteria for ‘low carbohydrate’. The characteristics of the 18 controlled trials are summarised in *Table 3*. The publication period covered 2006 to 2020, study duration ranged from 3 to 24 months, and study sample sizes ranged from 6 to 262 participants in the intervention arm. Of the included studies, 16 were randomised controlled trials (RCT) and 2 were non-RCT. Of the two non-RCT, one was randomised at the level of GP practice rather than participant level.

**Table 3:**
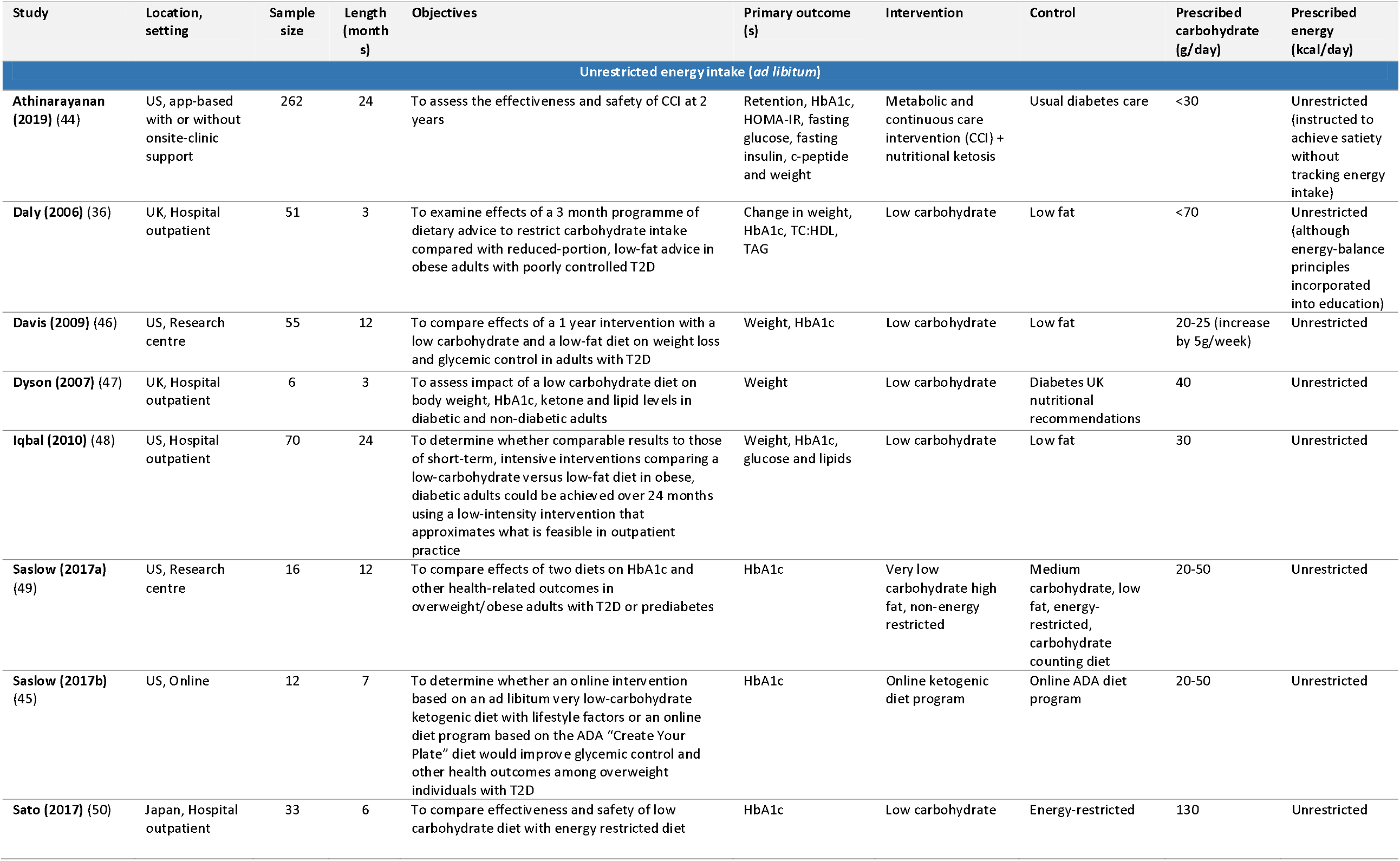
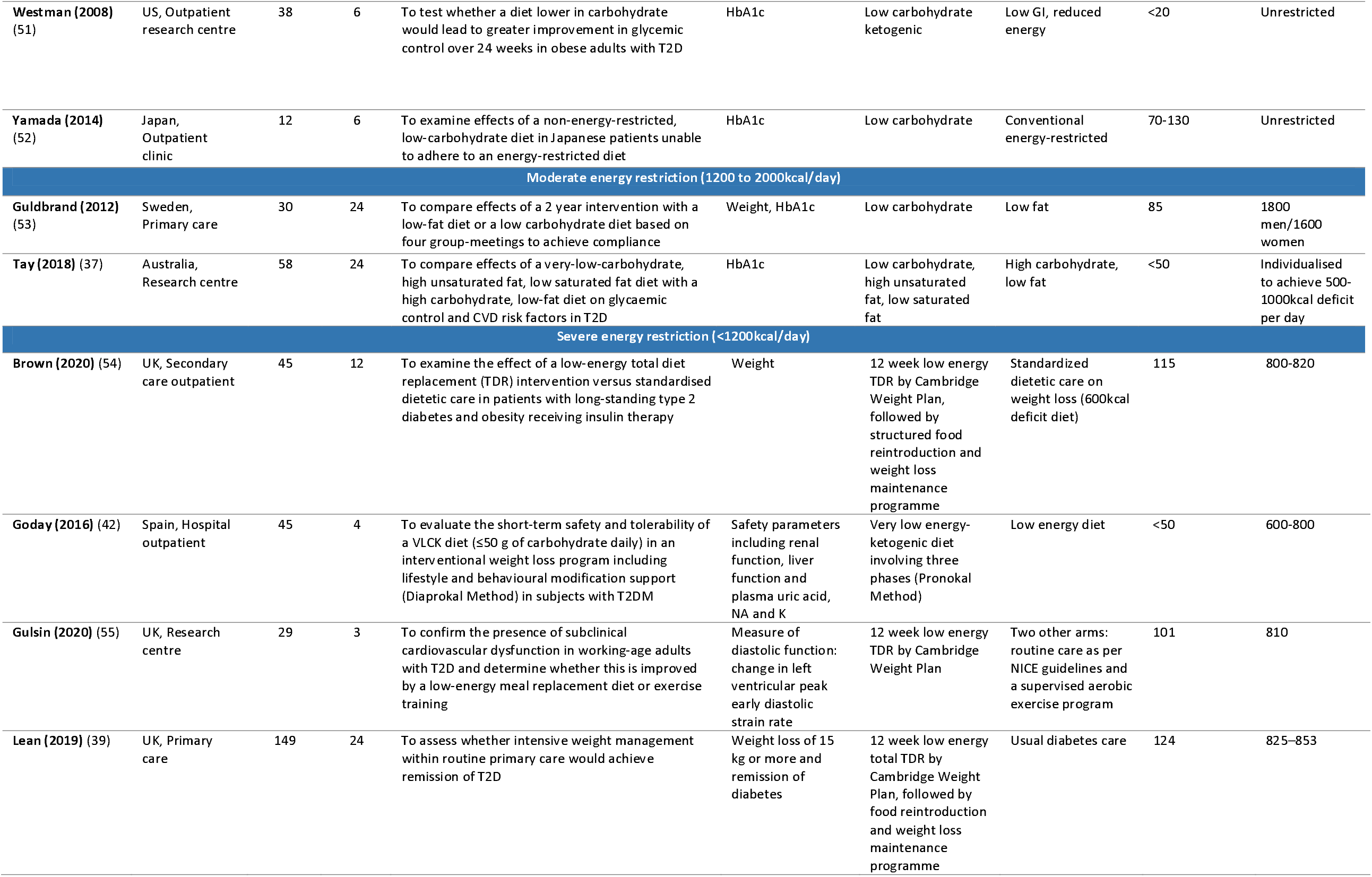
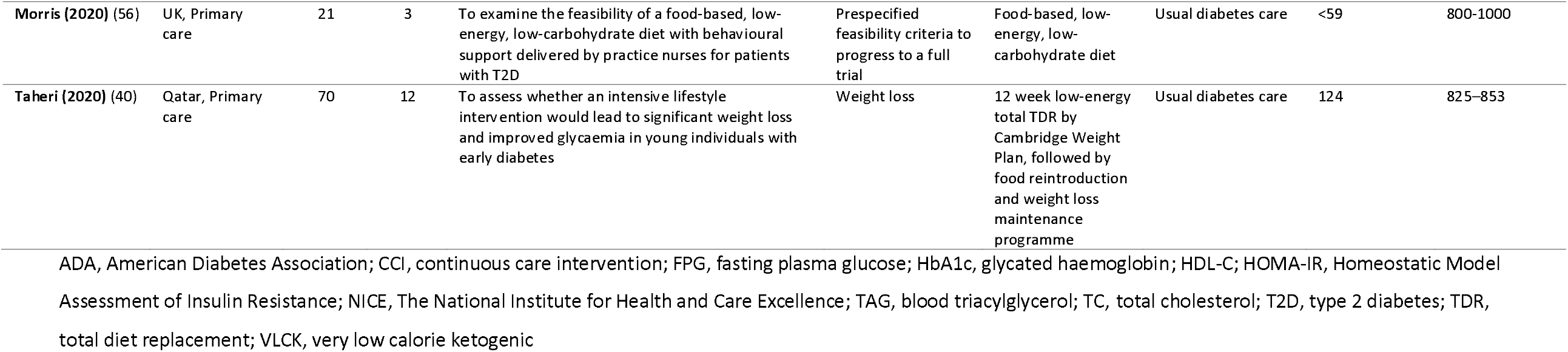
Characteristics of included studies.

### Variable approaches to energy and carbohydrate restriction

Studies were categorised into three groups based on their prescribed energy intakes: unrestricted (*ad libitum* feeding), moderately restricted (1200 to 2000 kcal/day) or severely restricted (<1200 kcal/day) (Figure 3).

**Figure 3:**
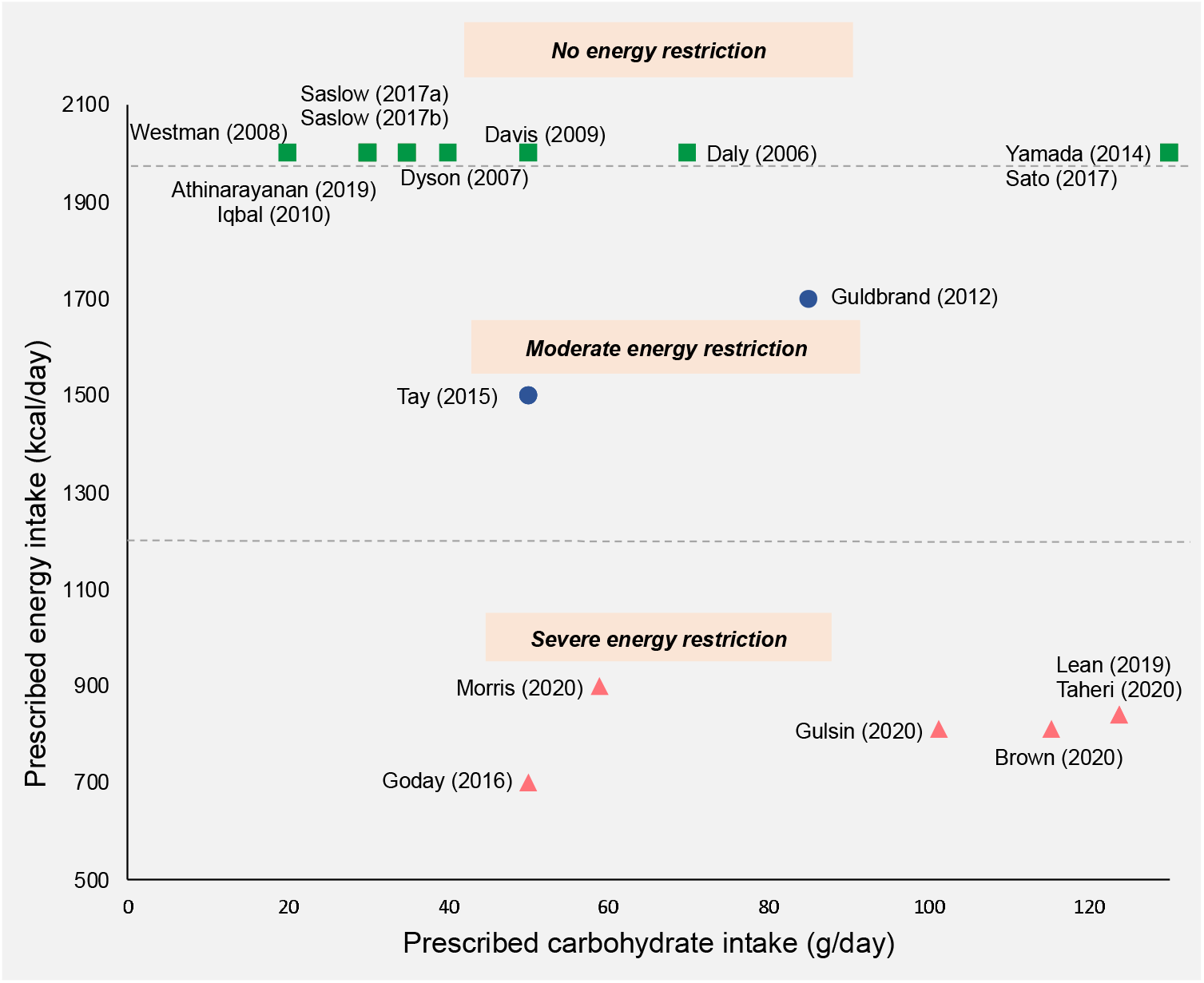
Prescribed daily carbohydrate and energy intakes in included studies. Where a maximum allowance of carbohydrate or energy was prescribed, this value was used. Where a range of carbohydrate or energy intakes was prescribed, the mid-point value was taken. Where energy intake was unrestricted, a value of 2000kcal/day was assigned. Squares, no energy restriction (ad libitum feeding); circles, moderate energy restriction (1200-2000kcal/day); triangles, severe energy-restriction (<1200kcal/day).

A total of 10 out of 18 studies involved traditional low or very low carbohydrate diets that were food-based and did not prescribe limits on energy intake. Two out of 18 studies used moderate energy reduction targets. Almost all of the studies in these groups aimed for sustained carbohydrate restriction throughout the study period. Only two were initiated by a very restrictive early phase followed by subsequent increases in carbohydrate allowance (36,37).

Six out of the 18 trials restricted energy intake to <1000kcal/day, with all except one incorporating meal-replacements as part of a stepped weight loss programme. Three studies used a 3 to 5 month proprietary total diet replacement (TDR) weight loss phase involving energy restriction of around 800kcal/day, followed by food reintroduction and weight loss maintenance phases (38–40). In the TDR phase, carbohydrates accounted for 50-59% of total energy (TE) which translated to 100-125g per day. The TDR phase was low carbohydrate in absolute terms but the macronutrient composition of subsequent phases was unclear from the published reports. These studies did not explicitly prescribe carbohydrate restriction but rather aimed to achieve weight loss via energy restriction. There were two exceptions to this; Morris (2020) (41) used a food-based diet that was explicitly low carbohydrate (<26% TE) as well as low energy (800-1000kcal/day) (39) and Goday (2016) (42) used a very low energy ketogenic diet. All of the LED trials were recently published, allowing only a limited period for follow up.

### Study aims

The principle aim for the majority of energy-restricted studies was to achieve and maintain weight loss, which was accomplished using mixed interventions with a number of different strategies including meal replacements to achieve <1200kcal/day, individualised dietary advice, lifestyle changes and medication, if necessary (43). For these studies, the control group was a variant of usual care. By contrast, the principal aim of most of the low carbohydrate diet studies was to test the effect of manipulating carbohydrate intake on participants’ weight and glycemic control, with a more conventional “low fat” diet as a comparator.

### Outcome measures

Most studies’ primary outcome measures related to weight and/or glycemic control with the exception of three studies that assessed cardiac function, safety parameters and full-trial feasibility criteria. All studies reported HbA1c, with 11 out of 18 studies specifying HbA1c as a primary outcome. Four studies, all published in or after 2018, reported T2D “remission” or “reversal”, although definitions of this varied (38–40,44). One earlier study reported the proportion of participants with final HbA1c below the T2D diagnostic criteria (45).

### Baseline participant characteristics

Mean population ages ranged from 42 to 69 years and there was a mix of ethnicities and gender ratios across studies. All studies except those conducted in Japan recruited participants who had BMI >30 kg/m^2^, with the majority having a mean BMI >35kg/m^2^. There was a wide range of average diabetes duration (2 to 14 years) and medication usage (Table S2).

### Intervention details

Interventions varied across studies in several ways including mode of delivery, dietary advice provided, intensity of support and utilisation of behavioural strategies to promote adherence (Table S3). Some studies included very low intensity interventions (involving only infrequent group sessions and dietary advice), whereas others involved more intensive input and employed a range of strategies to support dietary and lifestyle change including behaviour change techniques, intensive group support, biomarker feedback and health technologies, online support and remote care. More recent studies, published in or after 2017, involved higher intensity mixed interventions, typically employing a range of technological and behavioural support.

### Control

Six out of 18 studies used usual care as a control. In five of these studies, usual care provided minimal input so the intervention and control arms differed in a number of aspects beyond dietary change. The remaining 12 studies all used a version of a low fat, energy-restricted diet. In all but one study, energy intake was not matched between intervention and control diets. The exception was Tay (2018) who used a planned isocaloric control, advising both arms to limit energy intake to achieve a 500-1000kcal deficit per day.

### Dietary assessment and adherence

The majority of studies attempted to assess dietary adherence in some way, although the methods employed varied between studies. A total of 10 studies used food records (1, 3, 5 or 7-day food diaries) and two studies used self-report via questionnaires. Dietary adherence was not assessed in any of the four studies that utilised total diet replacement (via meal replacements). Several studies used urinary or blood ketones as a marker of nutritional ketosis but the majority of these did not base adherence on these reported measures. Of note, in those that included participant reported carbohydrate intakes, 8 out of 9 studies (90%) had reported values that exceeded the prescribed carbohydrate intake by more than 10% (Table S4).

### Risk of bias

Selection bias was high for the two studies in which participants self-selected into the intervention; it was low or unclear for the 16 trials that randomised participants between intervention arms. Six studies provided insufficient information on allocation concealment. Performance bias was judged to be high in the eight studies which involved mixed interventions that differed in several aspects to the control arm, due to the nature of these trials. Performance bias was unclear for the remainder of trials due to the possible influence of participant or personnel expectations on the results. Detection bias was considered low for all studies based on the objective nature of the outcomes of interest. Attrition bias was high in four studies in which dropout rates were high or imbalanced between groups and completers analysis was used. Reporting bias of the outcomes of interest was low in all studies since pre-specified outcomes of interest were reported. Only three studies were judged as unclear for ‘other bias’, due to potential baseline imbalances in confounders between the groups. The risk of bias assessment is summarised in Figure 4.

**Figure 4:**
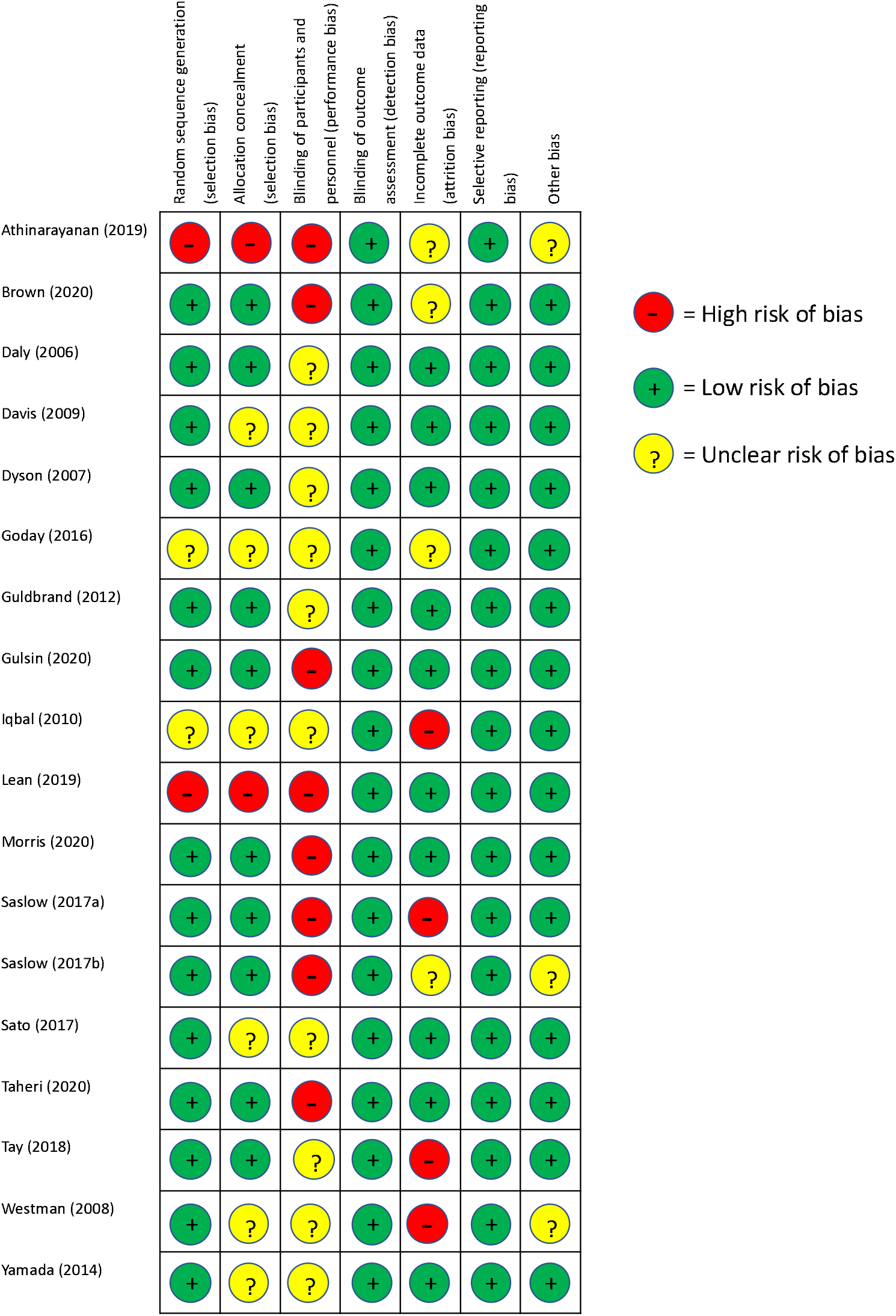
Risk of bias assessment results. +, low risk of bias; ?, unclear risk of bias; –, high risk of bias.

### Effectiveness of interventions

#### Between group differences

All but one study resulted in a reduction in HbA1c between baseline and study end-point (Table 4). A total of 10 studies demonstrated significant improvements in HbA1c in the intervention arm compared with the comparator arm. All studies reported weight loss from baseline to end-point, with 12 studies showing greater weight loss in the intervention arm compared with the control group. All studies using usual diabetes care as a control arm reported significant between-group differences in weight and HbA1c. Only two of the five studies reporting data at 24 months found a difference between intervention and control groups by the end of the study (39,44).

**Table 4:**
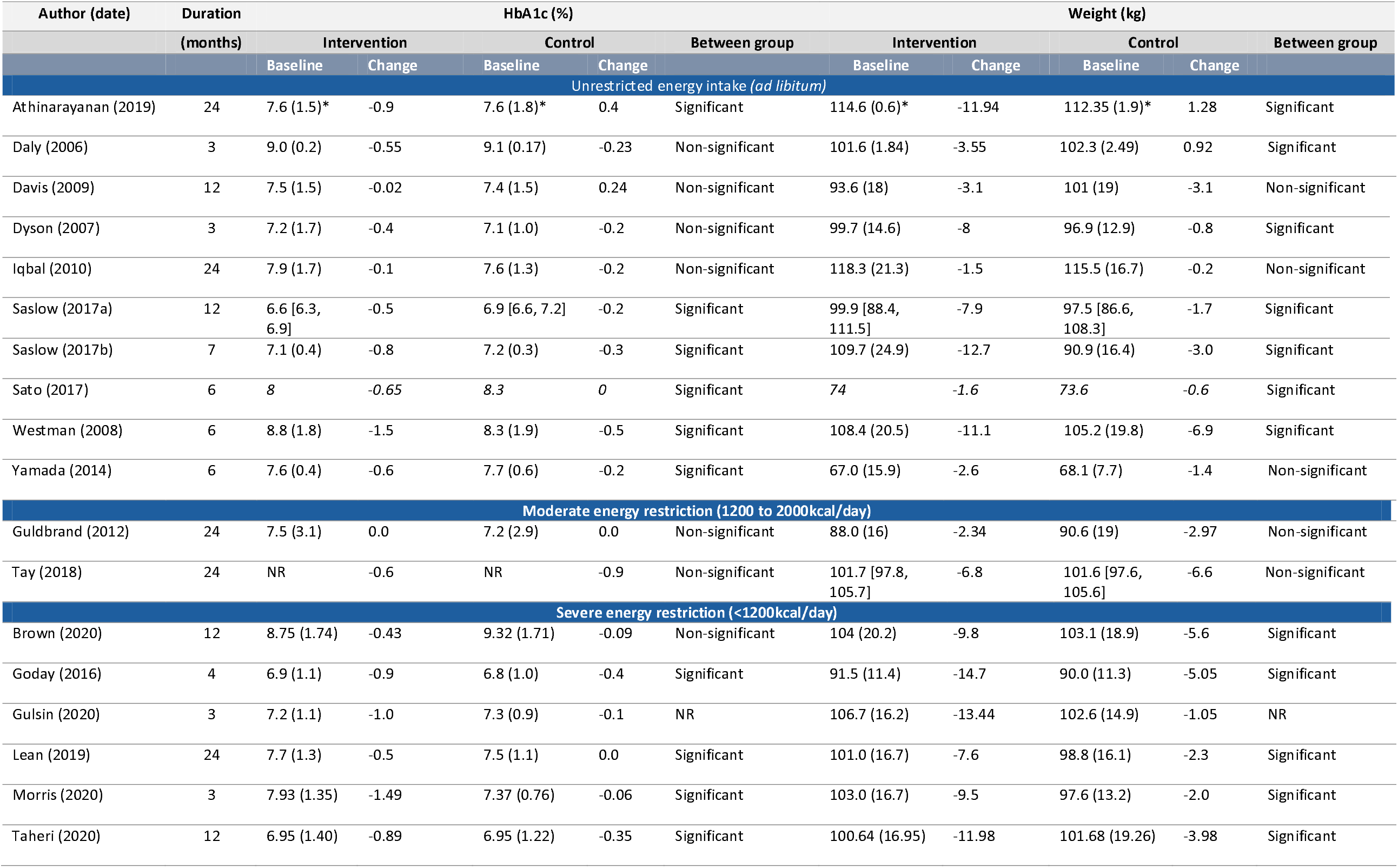
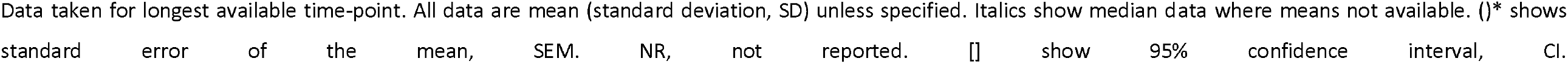
Baseline and change values for HbA1c and weight for intervention and control arms.

#### Weight loss and HbA1c change in intervention arms

There was a wide range of reported improvements in the intervention arms across studies in mean HbA1c change (ranging from 0.0% to 1.5%) and mean percentage weight loss (ranging from only 1% to over 15% of baseline weight). Figure 5 shows the data for all study endpoints.

**Figure 5:**
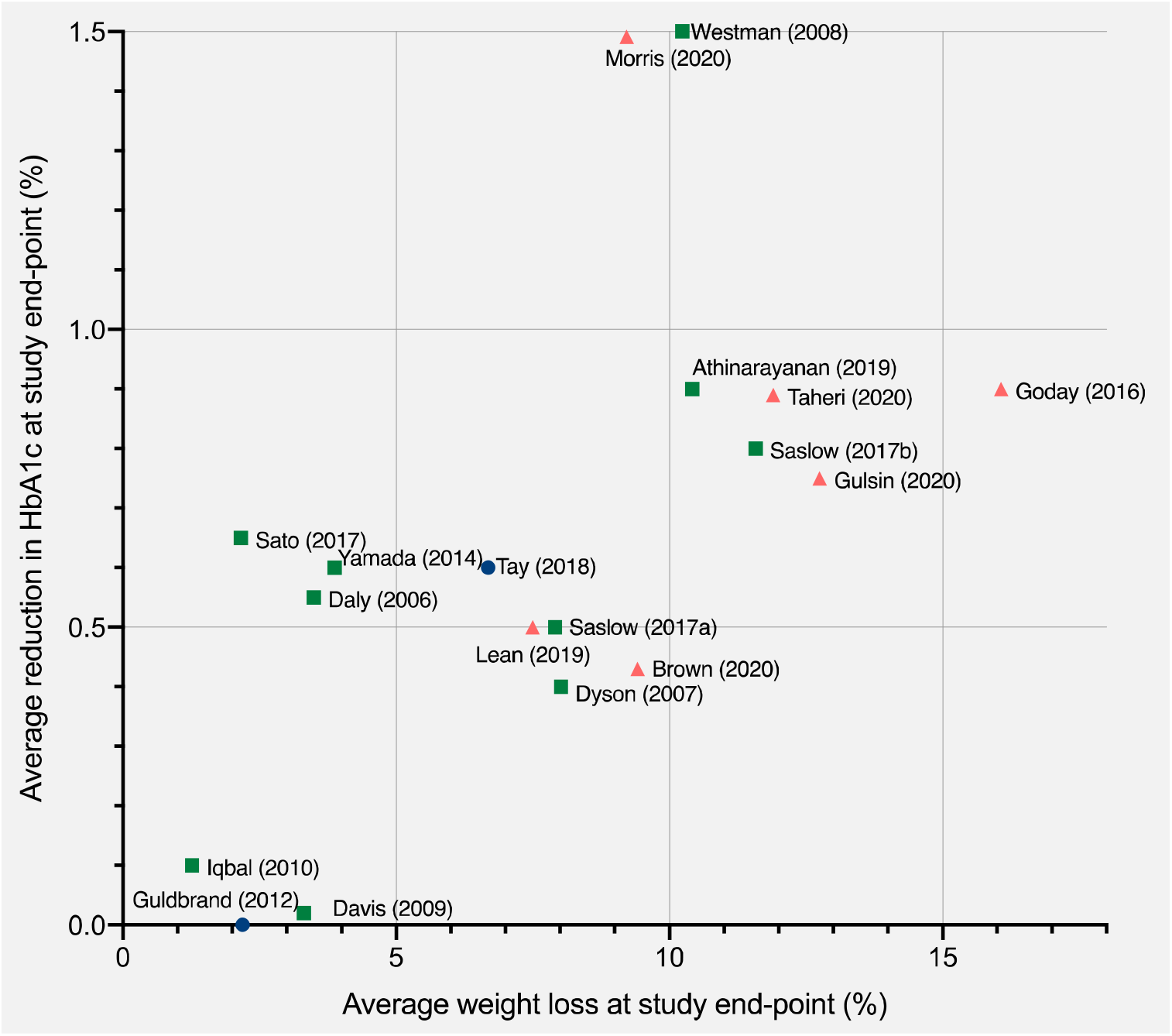
Average improvement in HbA1c and average percentage weight loss at study end-points. Each point represents the mean value for a single study with the exception of Sato (2017) which represents median values. Study end-points range from 3 to 24 months. Squares, no energy restriction (*ad libitum* feeding); circles, moderate energy restriction (1200-2000kcal/day); triangles, severe energy restriction (<1200kcal/day).

Those trials that severely restricted energy all produced clinically significant weight loss of more than 5% whereas energy-unrestricted trials produced a wider range of weight and HbA1c outcomes. The non-energy restricted studies were more numerous, published over a longer time period and involved more diverse intervention types. Two of the most effective interventions explicitly combined low carbohydrate and low energy approaches (41,42).

Figure 6 shows the data at 12 months to facilitate comparisons between studies. Level of energy restriction did not clearly distinguish intervention efficacy. The three studies using LEDs led to consistent and considerable mean weight loss of around 10% (16,39,40). The two studies reporting the largest changes at both 12 and 24 months involved LCDs with unrestricted or moderate energy restriction (37,44). The most effective intervention at 12 and 24 months involved an *ad libitum* ketogenic diet (44).

**Figure 6:**
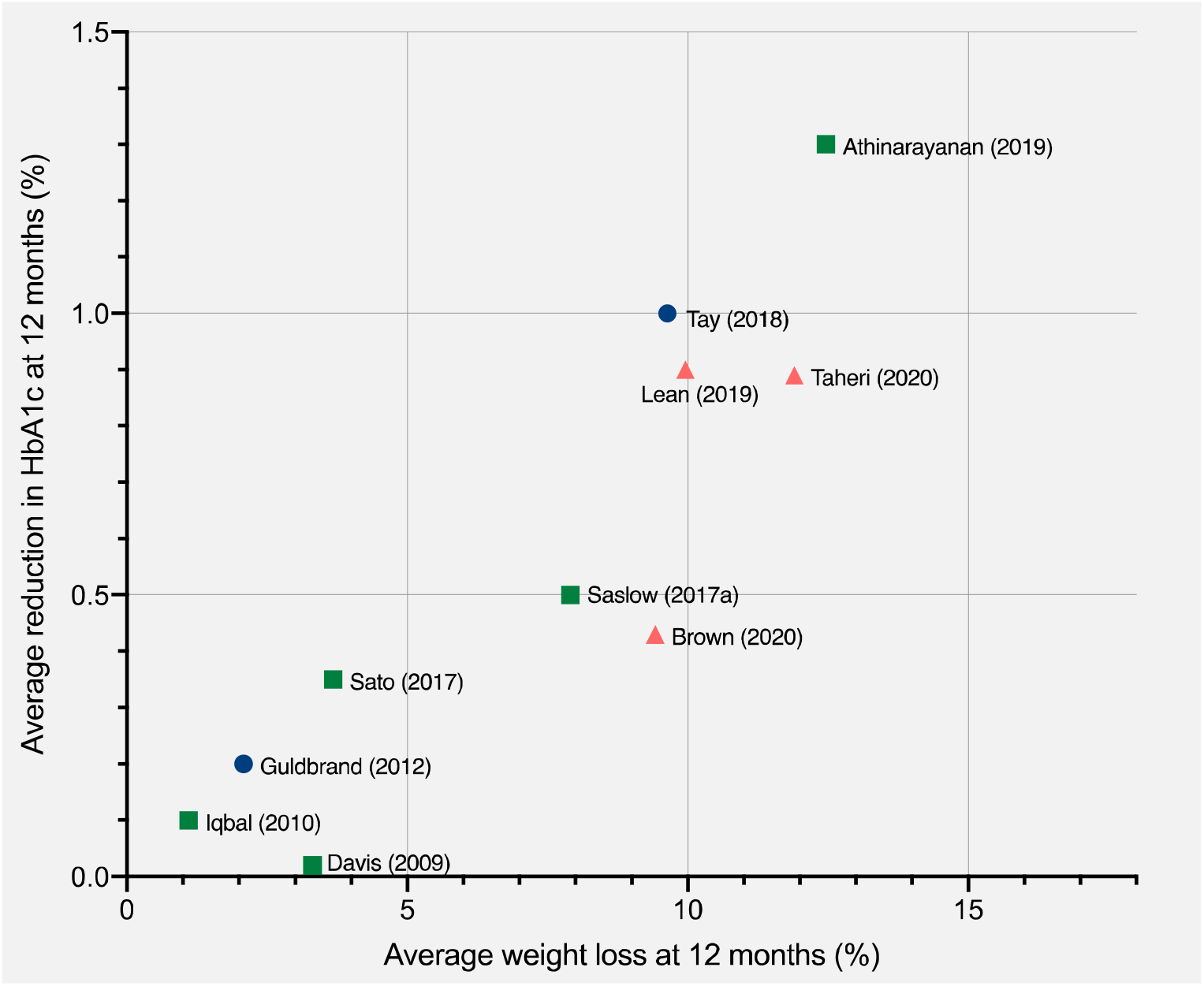
Average improvement in HbA1c and average percentage weight loss at 12 months. Each point represents the mean changes from baseline in HbA1c and weight for a single study, with the exception of Sato (2017) which represents median values. Studies were only included if they reported data at 12 months. Squares, no energy restriction (*ad libitum* feeding); circles, moderate energy restriction (1200-2000kcal/day); triangles, severe energy restriction (<1200kcal/day).

#### Association between weight loss and HbA1c

An association was observed between average weight loss and change in HbA1c across studies at 6, 12 and 24 months. The association was strongest at longer study-lengths, as assessed by R-squared: at 6 months 58% of the variation in average HbA1c change between studies could be accounted for by the variation in average weight loss; at 12 months this increased to 81%; and at 24 months this increased to 91%. A scatterplot summarises the results at 12 months (Figure 6). Reductions in HbA1c were associated with increased percentage of weight lost.

## Discussion

This systematic took a novel approach to the clinical trial evidence regarding dietary approaches to treat T2D by recognising that carbohydrate restriction is a common feature of LCDs and LEDs. Previous systematic reviews with meta-analyses have assessed the impact of higher versus lower carbohydrate diets (20–29). These have shown either no effect (27–29) or a positive effect of carbohydrate restriction on weight loss and HbA1c (20–26) and have noted the role of spontaneous energy restriction in LCDs as a potential confounder.

### Risk of bias

The heterogeneity of the study designs gave rise to different risk of bias when studies were evaluated with the Cochrane Risk of Bias tool. Studies that aimed to assess the efficacy of mixed interventions (involving dietary, physical, and behavioural changes) were judged as high risk of performance bias as they may document larger effects than those only assessing dietary changes. Additionally, two studies involved patients self-selecting the treatments they underwent which, although a valid approach for assessing efficacy, may also bias them towards reporting larger effects (39,44). The heterogeneity of the (mostly design-inherent) sources of bias precludes head-to-head comparisons.

### Intervention efficacy

This review found a range of intervention effectiveness that was not clearly distinguished by level of energy restriction: both energy-restricted and energy-unrestricted diets were effective at 12 months, and the most effective intervention at 12 and 24 months involved an *ad libitum* energy-unrestricted diet (44). This reinforces others’ observations of spontaneous energy restriction in LCDs (57) and highlights the potential efficacy of both low carbohydrate and low energy intervention types in the treatment of T2D.

The strength of the association between average weight loss and HbA1c change at 12 and 24 months was notable. This finding is consistent with the ‘Twin Cycle Hypothesis’ of T2D which proposes that T2D can be put into remission following weight loss, which reverses the accumulation of fat in the pancreatic beta-cells, thereby restoring their function (10). The potential causal relationship between weight loss and diabetes remission remains a matter of investigation (58,59).

Regardless of causality, the strength of the association between weight loss and glycemic markers underscores the importance of interventions that can maintain weight loss in the longer term. Weight maintenance is the most challenging area of weight management. Low energy meal-replacement based diets are capable of producing dramatic weight loss (60) but they are necessarily short term and weight regain is common upon cessation, especially in the absence of continued support (54). DiRECT (39) was the only LED trial to report data beyond 12 months and it will be of interest to see if the results achieved can be sustained over the full five-year trial period. This review identified a greater number of clinical trials testing low or very low carbohydrate diets, and a correspondingly wider range of outcomes. As with DiRECT, it will be of interest to see if the results using an *ad libitum* ketogenic diet in the Virta Health Study (Athinarayanan 2019) (44) can be maintained over the full five-year trial period.

In line with this focus on weight loss maintenance, this review identified a trend toward interventions with greater levels of participant support through co-interventions (involving exercise, pharmacotherapy, sleep and stress-reduction), new technologies and behaviour change techniques. Previous research shows that, regardless of modality of weight loss, participant support is important (61), and this represents a promising trend in research.

### Independent role of carbohydrate restriction

It is not clear from this review if carbohydrate restriction directly affects T2D status independent of weight loss. None of the included studies robustly measured energy intake or used an isoenergetic control, meaning the influence of spontaneous energy restriction was not controlled or accounted for. Tay (2018) (37) included a planned energy-matched high carbohydrate control but the diet was undertaken in a free-living environment and participants on the low carbohydrate arm reported lower energy intakes than those on the low fat diet. Several short-term studies do indicate a weight-independent effect of carbohydrate restriction on glycemic control (62–64) and there are other plausible underlying mechanisms that remain under investigation (65,66).

The field would greatly benefit from further research to explore the potential for an independent effect of carbohydrate-restriction on glycemic control. This could be tested using a parallel-arm clinical trial comparing low energy meal replacements with varying proportions of carbohydrates across a large enough range. Trials similar to this have been conducted using low energy formula diets with 100g (40%) versus 162.5g (65%) carbohydrates per day and 1000kcal for 4 weeks (67) and <40g versus 65-156g per day for 3 weeks each (in a crossover trial) (62). These trials have found that manipulating carbohydrates leads to differences in various markers of metabolic health. Trials using a broader range of carbohydrate intakes at fixed energy levels are needed to further explore these findings.

### Implications for clinical practice

The data in this review indicate that a major factor in T2D remission is weight loss maintenance. In clinical practice, patients would benefit from receiving information about the available options to enable them to make a fully informed individual choice, and to select for the diet and lifestyle changes that they can adhere to over the longer-term, which may or may not incorporate carbohydrate restriction.

### Limitations in the literature to date

The present review identified some limitations in the research to date. Inconsistent reporting of medication adjustment across studies means that changes in HbA1c were not considered in the context of medication changes. This can lead to an underestimation of the intervention impact on glycemic control (21,23,24,26,28). Future studies would benefit from a more standardised approach to reporting medication changes to facilitate comparisons between studies.

All of the included studies were conducted in free-living individuals, which enhances the ecological validity of the findings but reduces the level of control over participants’ diets. Diet studies in free-living individuals often suffer from poor adherence to the prescribed diet (68) and this review also found substantial deviations in reported versus prescribed carbohydrate levels among studies. For the majority of studies, participant intakes were assessed by self-report using food diaries or 24-hour recall. These methods have significant flaws which limit interpretation of achieved carbohydrate and energy intakes (69). Studies involving ketogenic diets may have an advantage since there is a measurable biomarker that can be used to assess adherence (44) but most trials did not report blood ketone levels.

During the selection and screening of studies for inclusion in this review, variation in definitions of ‘low carbohydrate’ were identified, something which is an ongoing challenge in this area of research. For consistency going forwards, the field would benefit from adhering to the definitions outlined by Feinman (18). The field would also benefit from standardisation of definitions of ‘low carbohydrate’ and ‘short/medium/long-term’ to ensure consistency in reporting.

### Limitations of present review

There are several limitations to this review. Firstly, presenting average weight and HbA1c outcomes of studies did not account for the underlying individual variability in weight and HbA1c outcomes. Secondly, intervention efficacy was based solely on weight and HbA1c change. Some studies reported outcomes including sleep quality, anxiety, and quality of life, as well as other glycemic outcomes such as fasting blood glucose and glycemic variability. There is also growing use and application of continuous glucose monitoring which provides measures of short-term glycemic control such as time in target range (70,71). Future reviews could consider inclusion of these and other outcomes to provide a more holistic review of the effectiveness of LED and LCDs in treatment of T2D. Thirdly, it did not distinguish between ketogenic and non-ketogenic diets. Ketones have been shown to directly lower hyperglycemia by suppressing hepatic glucose output (72,73). However, the role of ketosis in long-term weight loss is contentious due in part to poor adherence rates to ketogenic diets in some clinical trials (48). This is reflected in a recent systematic review that found that low carbohydrate diets were more effective than very low carbohydrate ketogenic ones, an effect which diminished when adherence was accounted for (26).

## Conclusions

This review took a novel approach to the dietary strategies for T2D remission by recognising the commonality of carbohydrate restriction between LED and LCDs. It found that trials that severely restricted energy intake were not superior to those that allowed *ad libitum* low carbohydrate feeding (no prescribed energy deficit) at longer study durations (12 and 24 months). However, the strong association between average weight loss and HbA1c change that strengthened with time indicates that successful interventions for T2D are those that enable sustained weight loss in the longer-term. Further studies that carefully match carbohydrate and/or energy intake between arms are needed to establish the independent roles of carbohydrate and energy restriction in T2D treatment.

## Data Availability

Data will be available for research purposes upon request to the corresponding author.

## Acknowledgments

None.

## Authorship

AN conceptualised the project, developed the search strategy, conducted the literature search, selected studies, extracted and interpreted the data, and wrote the manuscript. ASM contributed to the search strategy design, selected studies, reviewed extracted data and reviewed/edited the manuscript. HL and AC contributed to the project conceptualisation and search strategy design, provided final decisions on study selection and reviewed the manuscript.

